# Early Detection of Alzheimer’s Disease with Low-Cost Neuropsychological Tests: A Novel Predict-Diagnose Approach using Recurrent Neural Networks

**DOI:** 10.1101/2021.01.17.21249822

**Authors:** Devarshi Mukherji, Manibrata Mukherji, Nivedita Mukherji, Alzheimer’s Disease Neuroimaging Initiative

## Abstract

Alzheimer’s Disease (AD) is the most expensive and currently incurable disease that affects a large number of the elderly globally. One in five Medicare dollars is spent on AD-related tests and treatments. Accurate AD diagnosis is critical but often involves invasive and expensive tests that include brain scans and spinal taps. Recommending these tests for only patients who are likely to develop the disease will save families of cognitively normal individuals and hospitals from unnecessary expenditures. Moreover, many of the subjects chosen for clinical trials for AD therapies never develop any cognitive impairment and prove not to be ideal candidates for those trials. It is thereby critical to find inexpensive ways to first identify individuals who are likely to develop cognitive impairment and focus attention on them for in-depth testing, diagnosing, and clinical trial participation. Research shows that AD is a slowly progressing disease. This slow progression allows for early detection and treatment, but more importantly, gives the opportunity to predict the likelihood of disease development from early indications of memory lapses. Neuropsychological tests have been shown to be effective in identifying cognitive impairment. Relying exclusively on a set of longitudinal neuropsychological test data available from the ADNI database, this paper has developed Recurrent Neural Networks (RNN) to diagnose the current and predict the future cognitive states of individuals. The RNNs use sequence prediction techniques to predict test scores for two to four years in the future. The predicted scores and predictions of cognitive states based on them showed a high level of accuracy for a group of test subjects, when compared with their known future cognitive assessments conducted by ADNI. This shows that a battery of neuropsychological tests can be used to track the cognitive states of people above a certain age and identify those who are likely to develop cognitive impairment in the future. This ability to triage individuals into those who are likely to remain normal and those who will develop cognitive impairment in the future, advances the quest to find appropriate candidates for invasive tests like spinal taps for disease identification, and the ability to identify suitable candidates for clinical trials.

## 1 Introduction

Alzheimer’s Disease (AD) is a neurodegenerative disease that affects around 50 million people globally [17]. Projections show that 1 in 85 people will develop AD by 2050. In spite of the scale of the problem, there is no cure for the disease and countless clinical trials have been unsuccessful in finding effective treatment [5, 8]. In 2018, the Alzheimer’s Association estimated that the average cost for caring for an Alzheimer’s Disease patient is $350,174 – making it the most expensive disease in the United States.

Research shows that AD is an incredibly slowly progressing disease [3, 23, 9] and takes many years from initial cognitive decline to full-blown disease development. Due to this slow progression, early detection of AD can be crucial in both its treatment and prevention. Currently, the detection of AD relies on invasive and expensive tests - spinal taps for CSF Tau protein, brain scans, and blood biomarker detections [20, 12, 19]. These tests contribute to the overall costs associated with AD assessment and treatment mentioned above. It is therefore critical to develop an effective, less expensive, and unintrusive screening method to identify people who are at risk of developing AD so that the expensive and intrusive tests can be used only for those patients. Moreover, multiple ongoing global efforts aim to identify the optimal candidates for their clinical trials (cohorts) to test new therapies and understand disease progression. The cohorts selected for these trials often include patients who do not develop cognitive impairment during their participation and result in an unfortunate waste of resources [19, 7].

It has been shown that neuropsychological tests are effective in the diagnosis of AD and in the identification of patients that are likely to experience AD progression [22, 2, 18, 1, 4, 25, 11]. These tests are much cheaper to administer than CSF spinal taps and brain scans and can be used to identify patients at risk of developing cognitive decline and those that are not. The goal of this study is to use multiple Long-Short Term Memory Recurrent Neural Networks (LSTM RNNs) on a set of well-established neuropsychological tests such as the ones used by [24] to assess the current cognitive state of a subject and use current and past test score data to predict if a subject is likely to develop cognitive impairment within the next two to four years. This segmentation of subjects into those who are expected to remain cognitively normal and those likely to develop problems provides doctors and researchers an inexpensive and non-intrusive method to identify people that require more invasive interventions and also select appropriate subjects for clinical trials. Access to inexpensive assessments of cognitive state can be of great benefit to people around the world, especially those in underdeveloped countries where low-cost diagnosis methods can be used to preserve critical resources for only the most high-risk patients.

All data for this study were obtained from the Alzheimer’s Disease Neuroimaging Initiative (ADNI) database. ADNI was launched in 2003 in cooperation with the National Institutes of Health (NIH) and the National Institute on Aging to better understand AD development and discover new therapeutic techniques. This database includes data on neuropsychological tests, along with many other metrics used to monitor AD progression. Based on the research on neuropsychological tests in AD detection and prediction, five different neuropsychological tests (MMSE, ADAS Q4, ADAS Cog11, ADAS Cog13, and FAQ) were chosen for this study - [2] offers a brief description of these tests. The ADNI database includes data on all of these tests.

As mentioned before, this study uses Recurrent Neural Networks (RNNs) for diagnosis and prediction. RNNs are machine learning methods that use sequences of data to predict future values [15, 6, 10, 14]. This technique has been used effectively in many domains ranging from language recognition to marketing to healthcare. In the context of AD, [24] is one of the first studies that used RNN in AD prediction. Using NACC data, the paper used 78 features and a global CDR (Clinical Dementia Rating) score to identify patients that are likely to experience AD progression based on changes in CDR scores. Unlike ADNI, the NACC data are not collected at equal intervals for all subjects necessitating [24] to introduce time between visits as a separate feature. The paper utilized a large number of features (78) in a multi-feature model to identify patients that are likely to experience worsening of the disease in the future. In contrast, the RNNs used in the current study are developed individually for each of the five chosen tests and combined to diagnose a patient as likely to remain cognitively normal or not in the next two to four years. Consequently, this paper can be viewed as a step that should be conducted before a full-blown RNN that uses a comprehensive and expensive set of features is used to determine how rapidly a patient’s cognitive decline is expected to progress in the future. As noted earlier, it is not necessary to collect expensive data for patients that are unlikely to experience cognitive decline. The objectives of this study are:

1. To develop RNNs that use past and present neuropsychological test scores to diagnose whether a patient has cognitive impairment based on the values of each test.
2. To develop RNNs that can predict neuropsychological test scores over the next 2-4 years.
3. To combine the above two sets of RNNs to predict test scores for each test over the next 2-4 years and diagnose upon those predictions. These results determine whether a patient will stay cognitively normal in the next 2-4 years. Patients who are diagnosed as cognitively impaired or likely to experience impairment in the next 2-4 years will be recommended to undergo further assessment and treatment.

All of these objectives were met in this study.

## 2 Methods

### 2.1 Data

#### 2.1.1 Data, Software, and Packages

All data used in this project were obtained from the ADNI database. ADNI is a longitudinal study launched in 2003 in cooperation with the National Institutes of Health (NIH) and the National Institute on Aging. ADNI uses adult volunteers between the ages of 55 and 90. The initial cohort included 200 cognitively normal (CN) subjects, 400 subjects with mild cognitive impairment (MCI), and 200 subjects diagnosed with Alzheimer’s Disease (AD). Each individual is assigned a unique identification number or RID. CN subjects have a Clinical Dementia Rating (CDR) of 0 and MMSE score between 24 and 30; MCI subjects show signs of memory loss, have a CDR score of 0.5, and are at least one standard deviation below the mean score on the delayed recall portion of the Wechsler Memory Scale’s Logical Memory II. Those who are diagnosed with AD are diagnosed in accordance with the National Institute of Neurological and Communicative Disorders and Stroke-Alzheimer’s Disease and Related Disorders Association (NINCDS-ADRDA) [16]. The ADNI study has completed three phases to date and is currently in its ADNI3 phase. The first phase, ADNI1, started in October 2004 and spanned five years. ADNI-GO was the second phase and lasted two years after its launch in September 2009, and ADNI2 spanned five years since its launch in September 2011. In each phase, new participants were recruited, and participants from the previous phases continued. The ADNI dataset contains demographic data of the subjects as well as results of various neuropsychological tests, results of brain scans, and other physiological tests conducted at various intervals. The relevant data for this study are obtained from a subset of the ADNI dataset called ADNIMERGE and was downloaded on May 14, 2019. Data extraction and processing were conducted using the R programming language running in RStudio version 1.2. The recurrent neural network analysis was conducted using the Keras package version 2.2.5.0 running in RStudio. Descriptive statistical analysis was conducted using the statistical software package Stata 14.

Since the objectives of this paper are based on RNNs that use past and current neuropsychological test scores to diagnose a subject’s current cognitive state and use those values to predict future test scores and cognitive states, it is important to select subjects who have sufficient time series data for the chosen tests. A minimum of four data points is used for diagnosis and prediction. ADNI subjects undergo an initial assessment that determines their baseline cognitive states and then have follow-up tests at months 6, 12, 18, 24, 36, 48, 60,72, 84, and so on. The data for this study are primarily derived from the baseline, months 6, 12, 24, 36, 48, 60, and 72. (Months missing in this interval, such as 18, do not have any neuropsychological test values in the ADNI database.) A sequence of length four uses the baseline, and months 6, 12, and 24 data; a sequence of length six also includes months 36 and 48, and a sequence of length eight includes months 60 and 72.

#### 2.1.2 Neuropsychological Test Selections

Five neuropsychological test scores were used to assess the cognitive state of the subjects. The tests are MMSE, ADAS Q4, ADAS Cog-11, ADAS Cog-13, and FAQ. These tests have been used to demonstrate their effectiveness in diagnosis and prediction of cognitive impairment [2]. Additionally, neuropsychological tests were used in a regression model to determine the impact on change in CDR sum of scores [26] and “MMSE, FAQ, and ADAS-cog were identified as prognostic factors to detect cognitive decline in CDR-SB.” Other test scores, such as the Wechsler Logical Memory Delay (LDELTOTAL), are also available. However, the number of missing values of the latter test was greater than for the other tests, and the test was given less frequently (every two years as opposed to half-yearly or yearly) than the other tests, therefore limiting the number of subjects that could be used for the final test of the models, taking it out of consideration for this study.

#### 2.1.3 Data Imputation

As noted earlier, the battery of neuropsychological tests is administered during scheduled visits that occur at the initial visit (baseline) and then at months 6, 12, 24, and beyond. All participants do not attend all of the scheduled visits, and even when they do, results for all scheduled tests are not available. Consequently, there are many missing values for all of the tests that are part of the ADNI study. Moreover, since new subjects were added in the second and third phases of ADNI, not all subjects have data for all the month numbers listed above. Since the goal of this research is to start with at least four data points for a subject and predict the status in the next two to four years, sequences of length 4, 6, and 8 were extracted for each of the five neuropsychological tests separately from the ADNI1, ADNI-GO, and ADNI2 datasets and combined to form the training and validation data for our models.

Since the fundamental premise of this research is to learn from existing sequences of data for a specific test, project the trend into the future, and predict the state of disease progression at a future date, special care had to be taken to re-create or impute missing values. To use mostly real data values and reduce the number of imputations, subjects included in the training, validation, and test datasets had no more than 25% absences and missing test scores. Moreover, subjects who were absent or had a missing value on the last visit of a sequence of length 4, 6, or 8 were also dropped. The latter constraint ensured that no model would learn the generic trend of disease progression using imputed data at the most recent data point. These two steps resulted in an average of roughly 25% missingness in all the training and validations datasets.

A simple average method was used to impute missing values. As an example, for a model of length four, a subject with missing data for month 12 was assigned for month 12, the average of the baseline, month 6, and month 24 values. It is one of the many ways in which missing data can be imputed. This imputation method is advantageous since this method preserves the average of the sequence. The classification of a subject as cognitively normal or not is based on the average values of the sequences, as discussed below, and this imputation method replaces missing values without changing the averages in an arbitrary way.

### 2.2 Data Classification, Training, and Validation Set Creation

#### 2.2.1 Classification

Existing literature was used to determine the ranges of test scores for each of the tests noted above that can be used to distinguish people who are cognitively normal (CN) from those exhibiting cognitive impairment. Table 1 provides the range of test score values for each test, along with their cutoff scores for normal values. The last column of the table provides the normalization calculations used for each test to convert the data range to 0 to 1.

**Table 1:** Test Score Cutoffs and Normalizations.

| Test | Normal Cutoff | Min | Max | Normalization |
| --- | --- | --- | --- | --- |
| MMSE | $\geq 26$ | 0 | 30 | Value/30 |
| ADAS Q4 | $\leq 5$ | 0 | 10 | Value/10 |
| ADAS 11 | $\leq 10$ | 0 | 70 | Value/70 |
| ADAS 13 | $\leq 10$ | 0 | 85 | Value/85 |
| FAQ | $\leq 2$ | 0 | 30 | Value/30 |

To create the data for the diagnosis models, sequences of lengths 4, 6, and 8 from the combined ADNI1, ADNI-GO, and ADNI2 datasets were first separated into three different collections based on length. Then, each of the sequences in each collection was classified with either a 0 or a 1 according to the classification table criteria provided in Table 1. If the average of a sequence of data and the last data point in the sequence fell in the “Normal Test Values” range, it was assigned a 0 (CN). If the average and the last data point in the sequence fell outside of “Normal Test Values” range, the sequence was assigned a 1 (MCI/AD). Any sequence which did not satisfy the two latter constraints was dropped from consideration.

### 2.2.2 Diagnosis Model Data Creation

The resulting dataset was then sorted according to the classification value (0 or 1), and 80% of the smaller of the two classified sets was combined with a matching number of sequences from the other set of sequences to create the training set. This procedure was carefully incorporated due to the models’ enhanced ability to learn if there are an equal number of 0s and 1s. Finally, the remaining sequences were pulled together to form the validation data set.

Classifying the sequences allows for the diagnosis to be looked at from a neuropsychological test score standpoint. The efficacy of this novel technique is seen when the predicted diagnosis of this study is compared to the real diagnosis of these patients (seen in Results).

#### 2.2.3 Sequence Prediction Model Data Creation

For sequence prediction, the goal is to learn how to predict the data point t + 1 from data for sequence 1… t. The dataset with 6 value sequences combined from ADNI1, ADNIGO, and ADNI2 was used to generate training and validation data for the 5th and 6th value prediction models. For example, for the 5th value prediction model, the model was trained with sequence values 1 through 4 as input data and sequence value 5 as the output data value. The model learned how the 5th value was associated with the previous four values using the training data and tested what it learned on the validation data to determine its accuracy. Similarly, for the 6th value prediction model, sequence values 1 through 5 were used as the input data and value 6 was used as the output data value. Unlike in the diagnosis data creation, 80% of the rows classified as 0 were combined with 80% of the rows classified as 1 to create the training sets. The remaining 20% of rows were combined to create the validation sets. Rather than use an equal number of rows classified as 0s and 1s in the training sets, the models (in this case) performed best when there was as much data as possible to learn from. For the 7th and 8th element prediction models, a similar set of steps were followed using the combined data set containing sequences of length 8.

#### 2.2.4 Test Data Creation

To determine if a combination of neuropsychological tests can be used to predict the future cognitive state of an individual, it is necessary to create a test dataset of individuals for whom longitudinal data of length 8 (baseline to 72 months) are available for all of the five tests chosen for this study. The individuals’ test scores were predicted for the next two and four years and those predictions were collectively used to determine their future cognitive state. The same criterion of no greater than 25% missing values were used to select the subjects of the test dataset. As indicated before, many of the ADNI subjects do not have consistent data over a long duration for all of the relevant tests. To get subjects with long sequences of data, subjects were drawn exclusively from the oldest cohort - ADNI1. Care was also taken to ensure that these sequences were not used as training or validation data for any model. This process resulted in 66 distinct RIDs that met all the criteria for inclusion in the test dataset. Descriptive statistics for this cohort is given in Table 4.

### 2.3 Structure of the RNN Models

RNNs are neural networks that are able to process time-series data and make predictions upon that data. RNNs can be either used to predict categorical or quantitative outputs; only quantitative outputs were predicted in this study. The use of a basic RNN may lead to issues where the algorithm forgets data from the beginning of the sequence when handling longer time-series data. This dilemma is commonly referred to as the vanishing gradient problem due to the gradient-based learning approach of RNNs. A Long-Short Term Memory (LSTM) RNN uses a more complex set of layers (consisting of both tanh and sigmoid layers) inside each cell in the RNN to retain information when learning long time-series sequences. This results in an increased capacity for the model to remember information. Only LSTM RNNs were used in this study. The Keras package, which provides an R interface to the Keras neural network API, was used in this study to create the Recurrent Neural Network (RNN) models for diagnosis – the act of classifying a sequence of neuropsychological test scores as cognitively normal or abnormal.

#### 2.3.1 Model Creation

Each diagnosis and sequence prediction model was trained using its training data and validated using its validation data. The loss curve was plotted for each model, and both the model and the loss curve were saved. The models with the highest prediction accuracy were saved for later use in the test data prediction phase. The main goal and novelty of this study lies in the combination of the diagnostic and predictive models. The combination of the models is used on the final test data created for each neuropsychological test. The final test data set is the set of RIDs that is used to test the accuracy and practicality of the study, and it consists of 66 RIDs who all have test scores for each of the neuropsychological tests. The first four values of each of the sequences were fed into the best sequence prediction models, which predicted the 5th and 6th values of the sequence (two years into the future in relation to value 4). The best diagnosis prediction model was used to diagnose the predicted sequences of length six (diagnosis two years after value 4). The best sequence prediction model was then used again in order to predict the 7th and 8th values of each of the sequences. The best diagnosis prediction models were used to diagnose the predicted sequences of length eight (diagnosis four years after value 4). Then these diagnosis predictions, two and four years after the fourth value, were compared to the real diagnosis of the patients given by ADNI (using DXSUM values) at those respective times to determine the accuracy of the study as a whole.

## 3 Results

There are three sets of results for this work, and they relate to 1) accuracies of the diagnosis of cognitive states 2) accuracies of the predicted values of test scores two years (6th value) and four years (8th value) in advance, and 3) accuracies of the diagnosis and sequence prediction model combination on the test dataset. Table 2 gives the number of unique RIDs used to train and validate the CN/non-CN (0 or 1) diagnosis based on each of the five neuropsychological tests. The number of observations is sufficiently large for each test. The numbers show that for each test, the number of observations for the datasets decreases when going from sequences of length 4 to sequences of length 8 since the number of RIDs that have less than 25% missing values over a 72-month period or 48-month period is a lot less than it is over a 24-month period.

**Table 2:**
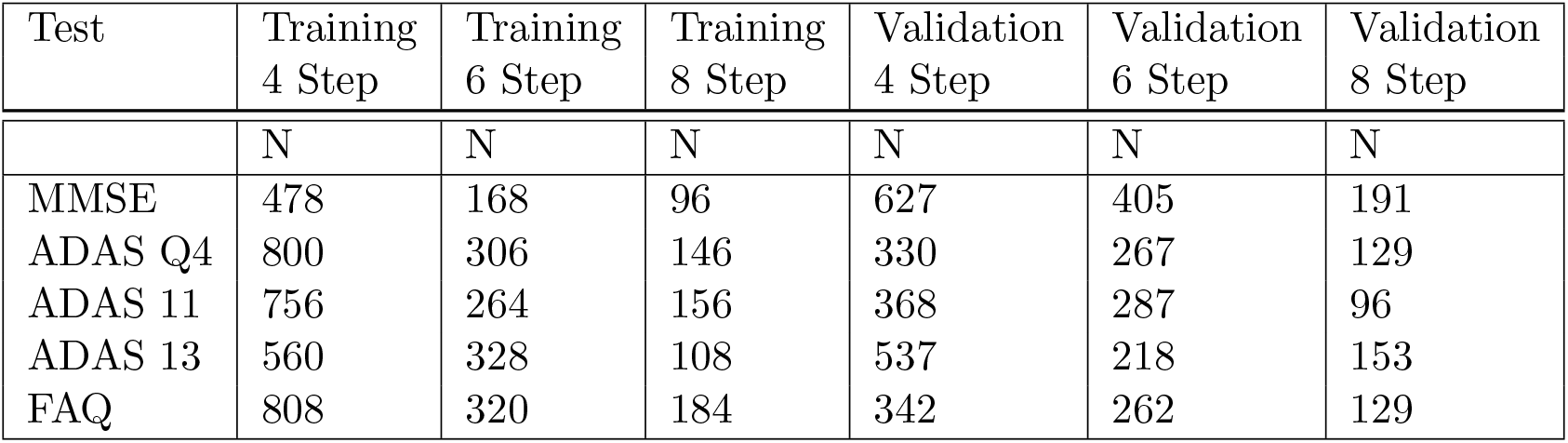
Number of Observations for Training and Validation Tests.

### 3.1 Diagnosis Models’ Accuracies

The diagnosis models learn from the training data to determine which sequences of validation data should be assigned a CN (0) value and which sequences should be assigned a non-CN (1) value. The models’ assignments of 0 and 1 are compared to the labeling of 0 and 1 of the validation data sequences during data classification. Table 3 gives the accuracies of the diagnosis models. The average accuracy is far above 90%. This shows that the models’ training have been effective in diagnosing subjects as cognitively normal or not. The accuracy values were determined by taking the proportion of correct diagnoses to the total number of diagnoses. The boxplots of Figure 1 serve as a corroboration of the accuracies noted above. In the classification technique, the sequences were classified based on the ranges provided in Table 1.

**Table 3:** Diagnosis Models’ Accuracies.

| Test | 4 Step | 6 Step | 8 Step |
| --- | --- | --- | --- |
| MMSE | 97.8% | 99.0% | 97.4% |
| ADAS Q4 | 98.8% | 99.6% | 96.9% |
| ADAS 11 | 91.0% | 97.9% | 96.9% |
| ADAS 13 | 97.4% | 97.2% | 97.4% |
| FAQ | 100.0% | 98.1% | 96.9% |

**Table 4.** Descriptive Statistics of 66 Test Dataset Subjects.

| Age | # | Gender | # | Education | # | Race | # | Marital Status | # |
| --- | --- | --- | --- | --- | --- | --- | --- | --- | --- |
| 58-68 | 8 | Female | 29 | 8-11 | 3 | Asian | 2 | Divorced | 4 |
| 68-78 | 41 | Male | 37 | 12-15 | 22 | Black | 4 | Married | 49 |
| 78-88 | 17 |  |  | 16-20 | 41 | White | 60 | Never married | 1 |
|  |  |  |  |  |  |  |  | Widowed | 12 |

**Figure 1:**
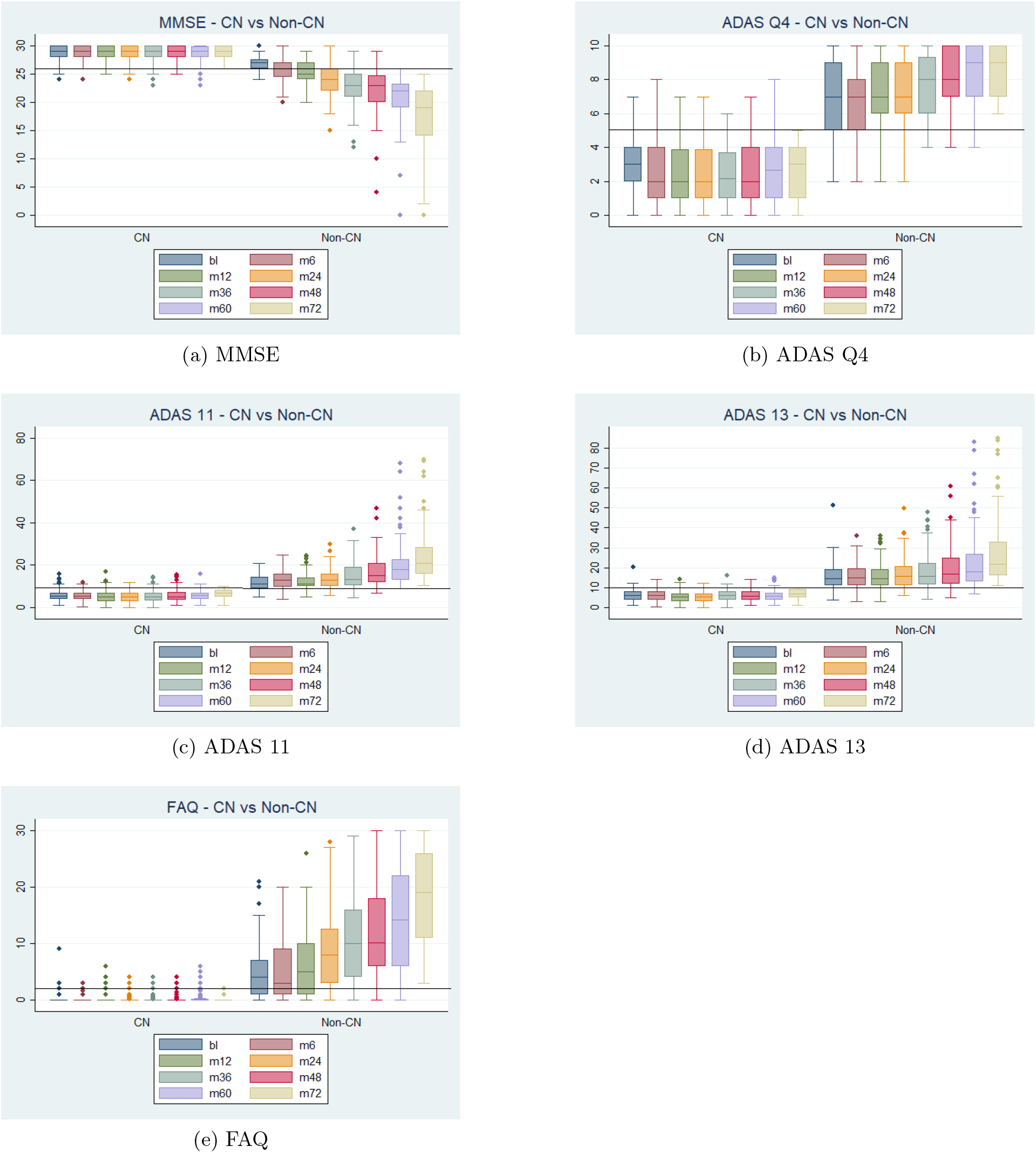
Ranges of Validation Data Split by the Predicted Diagnosis (CN or Non-CN) Assigned by the Diagnosis Models at Month 72 (with Normal Cutoffs)

The boxplots for each graph are split into two groups, CN and Non-CN. CN depicts the range of scores diagnosed as 0 by the diagnosis models, and Non-CN depicts those diagnosed as 1. The stark contrast in the ranges of the CN and Non-CN boxplots shows that the diagnosis models were able to effectively learn from the novel classification technique for all of the five tests.

### 3.2 Sequence Prediction Models’ Accuracies

Figure 2 consists of a series of graphs that demonstrate the accuracy of the sequence prediction models when predicting the 6th value based on the actual first four values and the predicted fifth value. The validation datasets were chosen for these figures. The blue line depicts the actual 6th value of each of the subjects, while the red line depicts the prediction of the 6th value using the best LSTM RNN model. The horizontal yellow line represents the threshold that differentiates test scores defined as CN or non-CN. The comparisons show that even if the predicted values are not exactly identical to the actual values, they are close and more importantly, following the general trend of the actual data. If the predicted values did not follow the general trend, the diagnoses based on these values would not be accurate. Moreover, the inaccuracy is of greater concern if the predicted and actual values lied on two different sides of the cutoff line.

**Figure 2:**
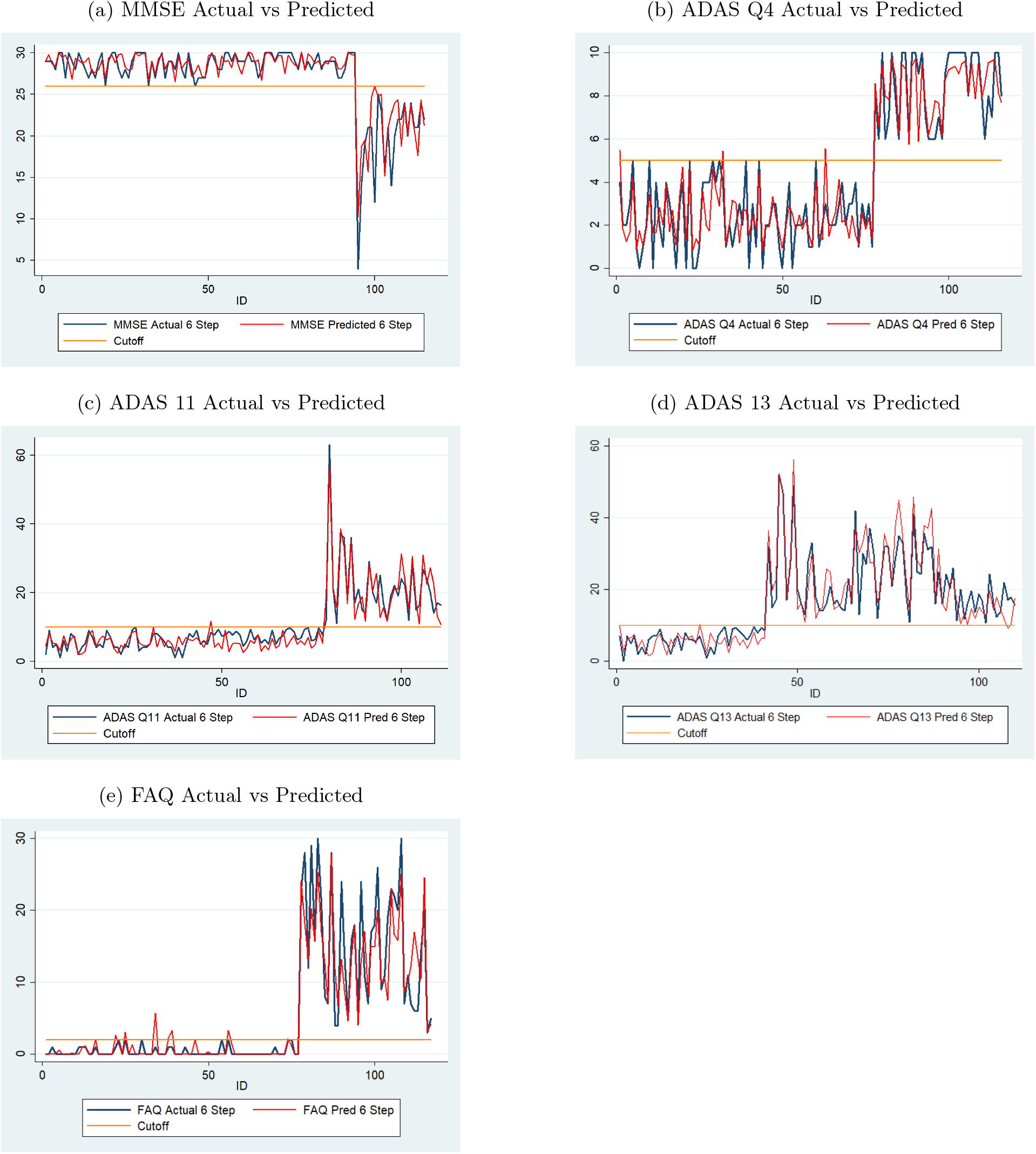
Comparison of Actual and Predicted Values by Sequence Prediction Models at Month 48 for Validation Data (with Normal Cutoffs)

Figure 3 displays the accuracy of the sequence prediction models when predicting the 8th value, the 72nd month, based on the actual first four values and the predicted fifth, sixth, and seventh values. In this case as well, the data are for the individuals in the validation datasets. The results are similar to those reported above for predicting the 6th value. Figures 4 and 5 display results similar to those in figures 2 and 3 with the exception that they are for the 66 individuals who are part of the test dataset. The results depicted in Figure 4 for predictions two years ahead, show that the percentage of predicted scores on the same side of the cutoff lines are high for all tests for month 48. The same is true for month 72 as shown in Figure 5. The percentages for month 72 are lower than the percentages for month 48 and is to be expected due to the fewer number of test subjects available for longer sequences.

**Figure 3:**
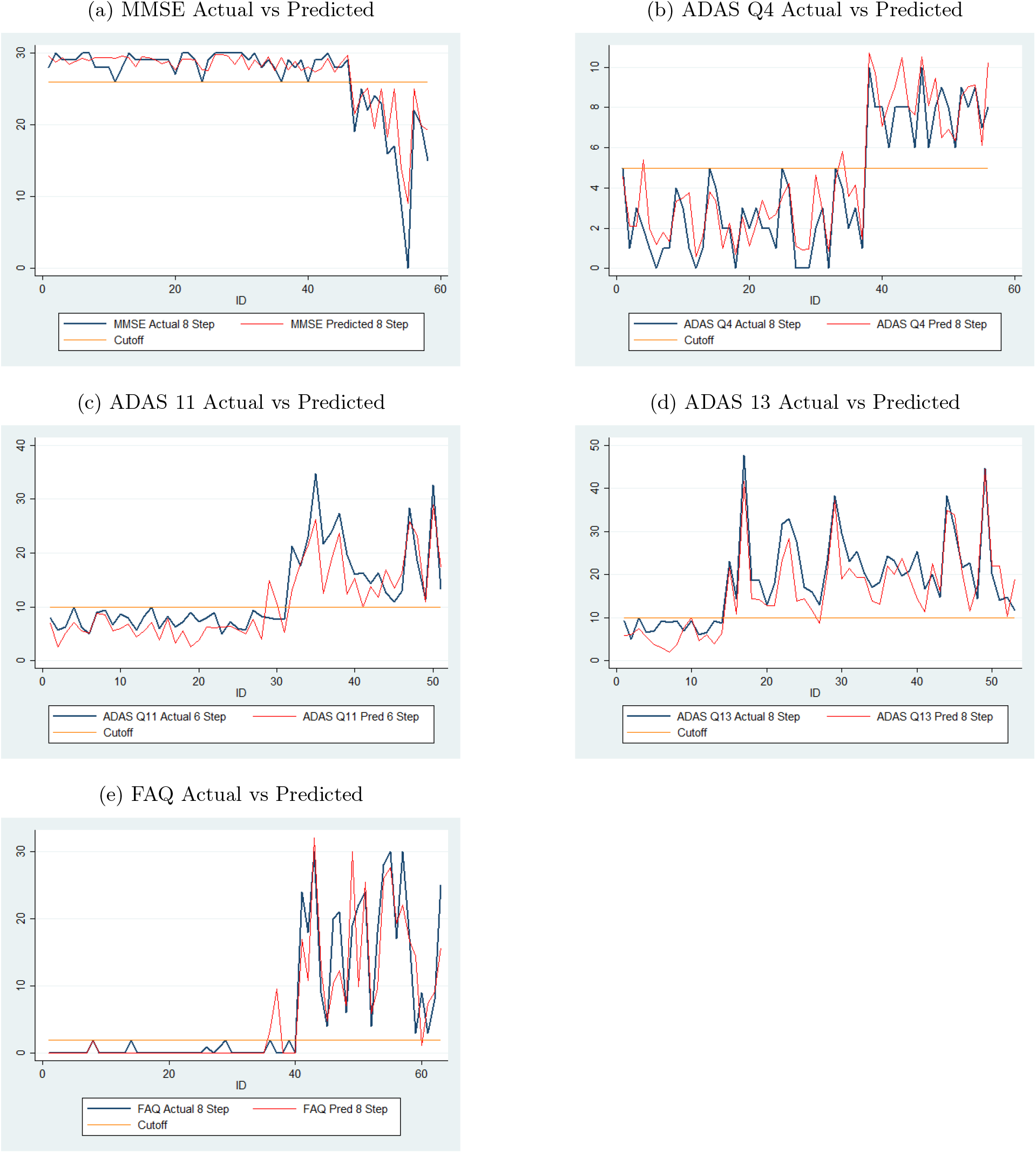
Comparison of Actual and Predicted Values by Sequence Prediction Models at Month 72 for Validation Data (with Normal Cutoffs)

**Figure 4:**
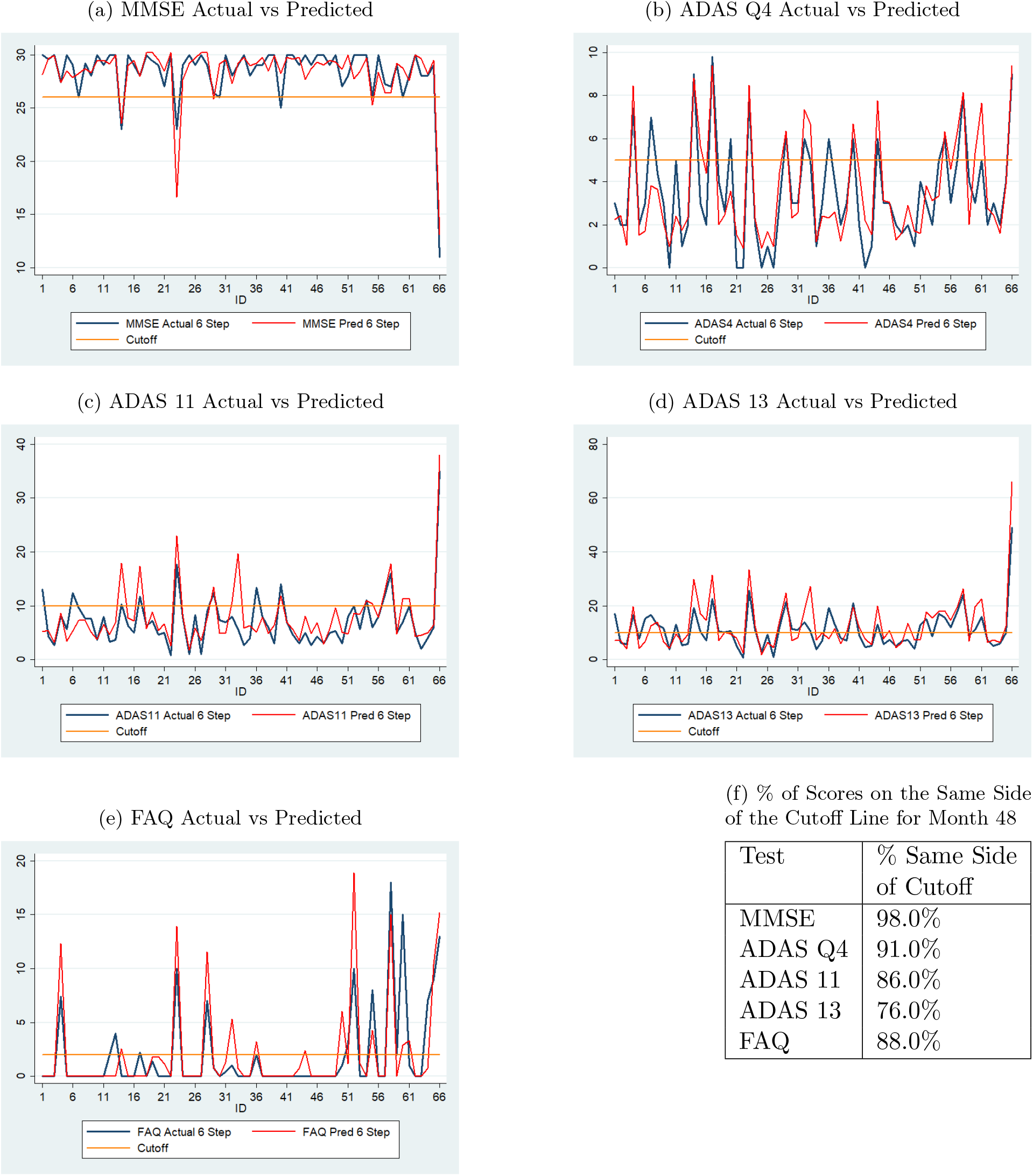
Comparison of Actual and Predicted Values by Sequence Prediction Models at Month 48 for 66 Test Dataset Subjects (with Normal Cutoffs)

**Figure 5:**
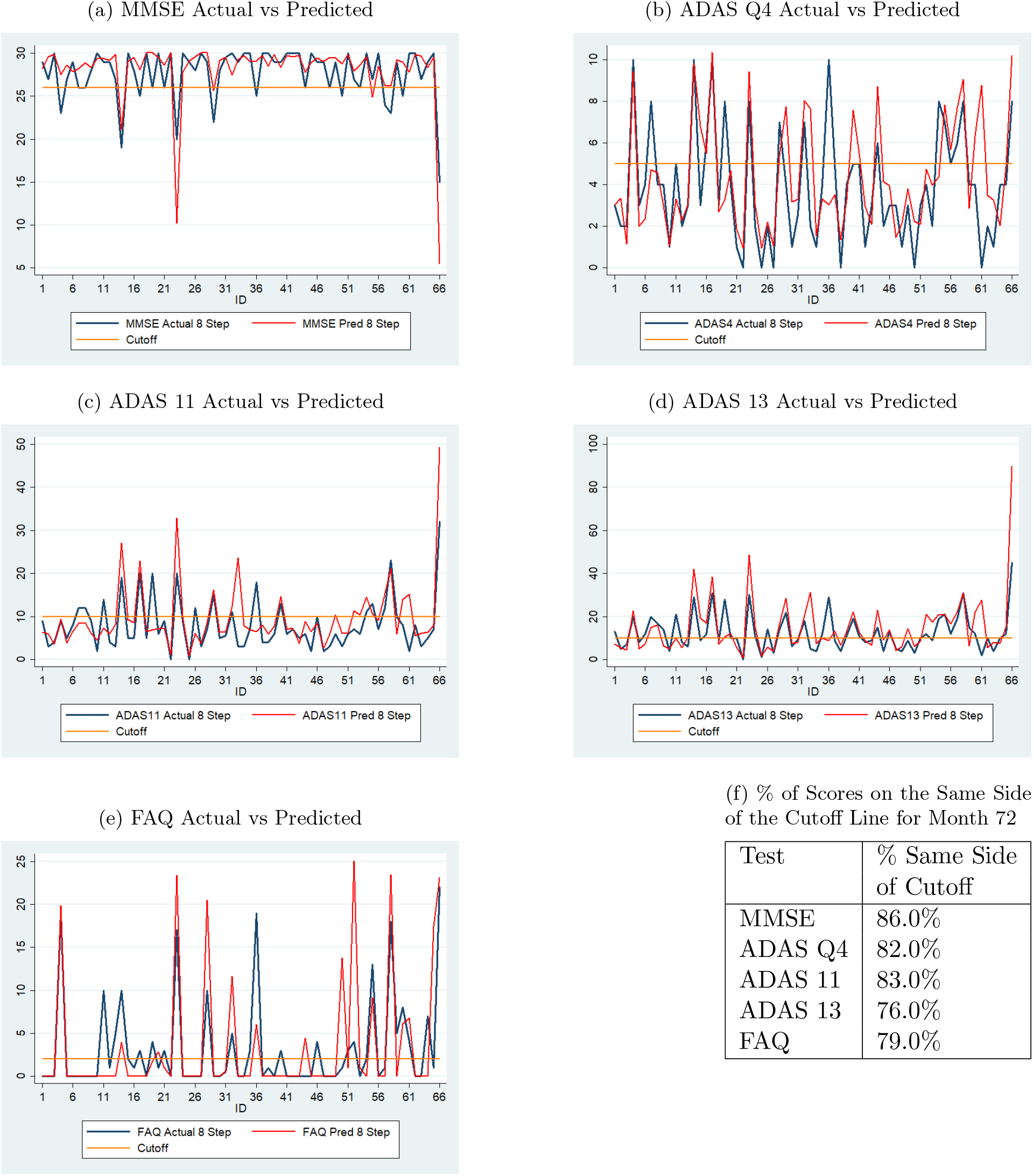
Comparison of Actual and Predicted Values by Sequence Prediction Models at Month 72 for 66 Test Dataset Subjects (with Normal Cutoffs)

### 3.3 Combined Model’s Accuracy

The key objective of this study is to determine if neuropsychological test scores can be used to predict if an individual will remain cognitively normal in the next two to four years. If the models, when combined together as illustrated in section 2.3.1, predict that a patient will not remain CN over the next two to four years, then those individuals should be monitored more frequently and are likely to be recommended to undergo further testing.

A relatively simple rule that is used to determine if a subject is expected to remain CN in the next two to four years is if their predicted 48-month and 72-month average test scores are in the normal ranges for all of the five neuropsychological tests used in this study. Tables 5 and 6 show the number of tests the 66 subjects are predicted to fail at months 48 and 72 (2 and 4 years after the fourth value, respectively) and the comparison to their actual diagnoses by ADNI as CN, MCI, or AD at months 48 and 72.

**Table 5a.**
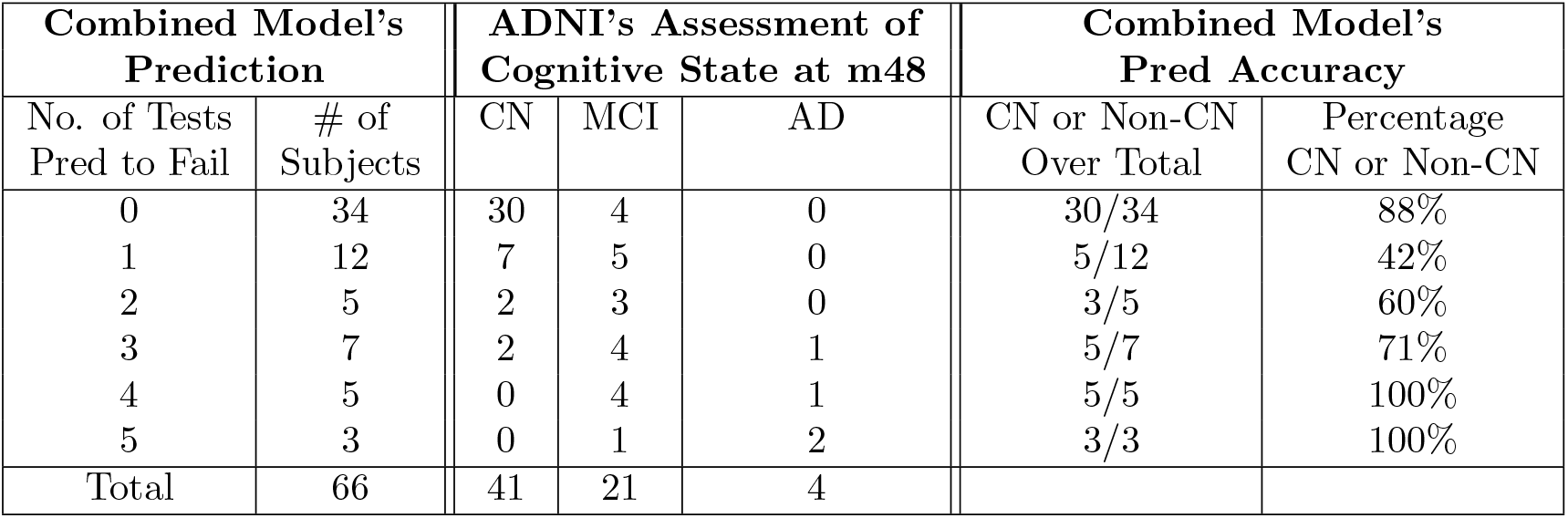
Combined Model’s Prediction of Number of Neuropsychological Tests Expected to Fail by Test Dataset Subjects at Month 48.

**Table 6a.**
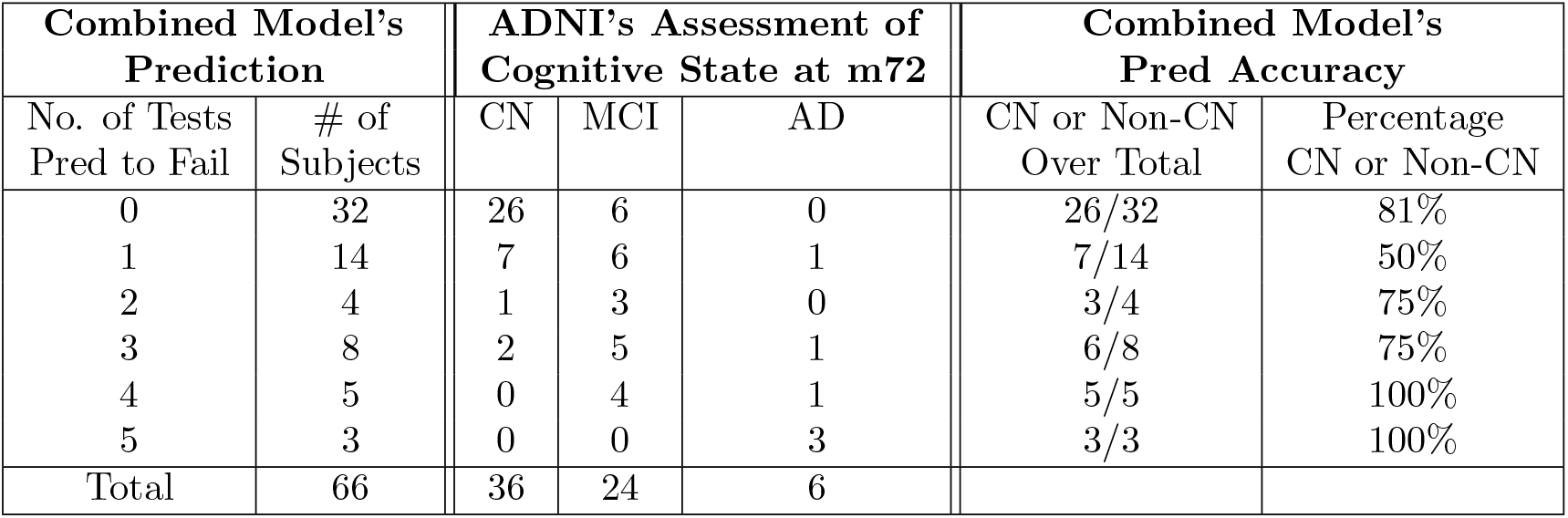
Combined Model’s Prediction of Number of Neuropsychological Tests Expected to Fail by Test Dataset Subjects at Month 72.

Table 5 shows that out of the 66 subjects in the test dataset, 34 subjects are predicted by the combined model to be fail 0 tests at month 48. Thirty of these 34 were diagnosed as CN by ADNI at month 48. Thus, 88% of the subjects remained CN as predicted by the combined model. The remaining four subjects are predicted by the model to pass all of the tests at months 48 but were classified by ADNI as MCI.

**Table 5b.** Combined Model’s Prediction Accuracy of Diagnosis at Month 48.

| Combined Model's Predictions |  |  | ADNI's Assessment |  | Accuracy |
| --- | --- | --- | --- | --- | --- |
| Tests Pred to Fail | Diagnosis | # of Subjects | CN | Non-CN |  |
| 0 | CN | 34 | 30 | 4 | **88%** (30/34) |
| 1 or More | Non-CN | 32 | 11 | 21 | **66%** (21/32) |

The 32 remaining test subjects were predicted to fail at least one test and would be recommended for more monitoring. Twelve subjects are predicted by the combined model to fail one of the five tests. Twenty subjects are predicted by the model to fail two or more tests at month 48. Table 5 shows that the percentage of subjects diagnosed as MCI/AD (non-CN) by ADNI increases from 42% for one test to 100% for four or more tests. 80% (16 out of 20) of the subjects the model predicted would fail two or more tests were diagnosed as MCI or AD by ADNI.

Table 5’s results for the prediction of cognitive state at month 48 based on actual test scores up to month 24 and predicted test scores for months 36 and 48 can be summarized as follows.

- 88% of the subjects predicted to remain cognitively normal two years in the future actually were diagnosed as cognitively normal by ADNI after two years.
- 80% of the subjects who were predicted to fail two or more tests two years in the future were diagnosed as cognitively abnormal (either MCI or AD) by ADNI.
- 42% of subjects who were predicted to fail only one test were diagnosed as cognitively abnormal (either MCI or AD) two years later.

These results show that subjects who are predicted to pass all tests in the next two years are highly likely to remain cognitively normal. In all other cases, further testing and monitoring are recommended.

Table 6 reports predictions made by the model for the number of tests the 66 test subjects are likely to fail four years later (at month 72) and compares them with actual diagnosis by ADNI at month 72. 32 of the 66 subjects are predicted by the model to pass all five tests at month 72. ADNI diagnosed 26 of them (or 81%) to be CN at month 72. 14 out of the rest are predicted to fail one test of whom 50% were diagnosed as non-CN by ADNI. 85% (17 out of 20) of the subjects predicted by the model to fail two or more tests are diagnosed as MCI or AD by ADNI at month 72. Table 6’s results are summarized as follows.

**Table 6b.**
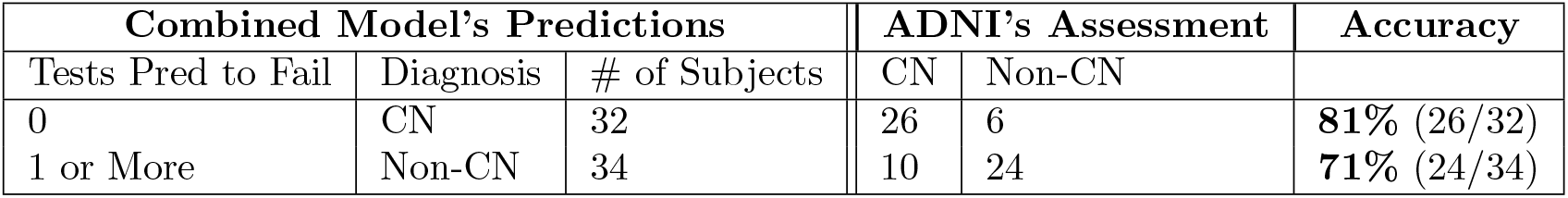
Combined Model’s Prediction Accuracy of Diagnosis at Month 72.

| Combined Model's Predictions |  |  | ADNI's Assessment |  | Accuracy |
| --- | --- | --- | --- | --- | --- |
| Tests Pred to Fail | Diagnosis | # of Subjects | CN | Non-CN |  |
| 0 | CN | 32 | 26 | 6 | **81%** (26/32) |
| 1 or More | Non-CN | 34 | 10 | 24 | **71%** (24/34) |

- 81% of the subjects predicted by the model to pass all tests at month 72, were diagnosed as CN at month 72 by ADNI.
- 50% of the subjects predicted to fail only one test were diagnosed as CN at month 72 by ADNI.
- 85% of subjects predicted to fail two or more tests, were diagnosed as cognitively abnormal (either MCI or AD) at month 72 by ADNI.

Tables 5 and 6 collectively show that the combined model is successful in predicting that an individual will remain cognitively normal in the next two years with 88% accuracy. This accuracy drops to 81% for predictions that are four years in the future. As mentioned earlier, the drop in accuracy is expected due to decreasing amounts of valid data as sequence length increases. In all other cases, the model finds a moderate to a high likelihood of the subjects experiencing cognitive challenges. These subjects are good candidates for more invasive testing and are likely to serve as good candidates for clinical trials for AD treatments.

## 4 Discussion and Conclusion

This study has demonstrated that it is possible to use machine learning tools on past and current neuropsychological test scores to predict future values of those scores and predict future cognitive state. Using data from hundreds of subjects who participated in the ADNI project, this study used RNN techniques to both diagnose individuals as CN or non-CN and predict future values of their tests scores. For a cohort of 66 test subjects, the test scores of all five tests were combined to generate a prediction of their future cognitive states based on the combined results. These results were then matched with the actual CN or MCI/AD status assigned by ADNI at future points in time. The results show that individuals who are predicted by the model to continue to remain CN in all of the tests are highly likely to remain cognitively normal in real life over the next two to four years. Individuals who are predicted to have test scores outside of normal ranges are likely to experience cognitive impairment in the future. These individuals should be monitored and treated.

Tables 5 and 6 show that for 66 test subjects, over 80% of those who are predicted to have all neuropsychological test scores in normal ranges are actually diagnosed as cognitively normal by ADNI at months 48 and 72. Over 80% of those predicted to fail 2 or more tests are likely to experience cognitive impairment. The results are more evenly split for subjects predicted to fail only one test.

The lack of accuracy related to failure of one test contributed significantly to the decline in overall accuracy of non-CN predictions. Upon further analysis it is revealed that for the month 48 prediction, 10 out of 12 subjects who are predicted to fail only one test, are expected to fail the ADAS 13 test. 70% of them were diagnosed as CN by ADNI at month 48. The other three were diagnosed as MCI. This predominance of ADAS 13 as the test individuals are likely to fail in case they are predicted to fail only one test is true for the month 72 prediction as well. These observations suggest that failing only ADAS 13 may not indicate the likelihood of developing serious cognitive issues within the next two years. It is also possible that this is an artifact of the cutoff score of 10 chosen for the ADAS 13 test to classify individuals as CN or non-CN. Test score data used for model training show that for the 328 individuals used for training purposes, ADAS 13 scores were below 10 for 75% of the individuals who were diagnosed as CN by ADNI. This suggests that the cutoff of 10 for CN subjects is an acceptable choice. [13] shows that the average ADAS 13 score for CN individuals is 9.24. Nonetheless, further analysis of the ADAS 13 test score as the only indicator of potential cognitive challenges is warranted particularly when the scores are close to the cutoff value of 10 (all of the 10 subjects predicted to fail only ADAS 13 had scores very close to the cutoff - between 10 and 13).

In addition to predicting if an individual is likely to remain in the cognitively normal range in the future, [21] study the predictive progression of cognitive impairment in MCI subjects in the future. While this progression or conversion is not the primary focus of this study, the boxplots in Figure 1 clearly show that individuals diagnosed as non-CN based on the cutoff scores chosen experienced significant worsening of their test scores over time. The boxes moved further away from the cutoff scores from the baseline to month 72 for all of the scores considered. In contrast, individuals diagnosed as CN had test scores that varied little from baseline to month 72.

The analysis of this paper focused on five tests for which consistent data over at least a 72 month period was available for a large number of individuals. Studies such as [2] used a battery of tests to determine which ones are most effective in classifying patients accurately with small differences in CDR scores. Their Figure 2 shows LDELTOTAL, LIMMTOTAL, Q4, TOTALMOD, Q1, MMSESCORE, TOTAL11, FAQTOTAL, and CATVEGESC to be some of the effective tests. This study used Q4 (ADAS Q4), TOTALMOD (ADAS 13), MMSE, TOTAL11 (ADAS 11), and FAQ from this list. The rest of the tests did not have sufficient longitudinal data to be used. Another study, [21], assessed different neuropsychological tests to determine which have good prediction power. While many of the tests identified by these studies had some data available in ADNI, the limitation of having no more than 25% missing values restricted the number of tests that could be considered for this study. The combined model’s structure allows one to introduce any test for which sufficient longitudinal data is available from ADNI or any other data source such as NACC (National Alzheimer’s Coordinating Center).

The benefit of developing a tool based on neuropsychological test scores that can predict with high accuracy the likelihood of an individual experiencing cognitive difficulties in the next few years derives from the low cost of administering these tests. If these tests become routine for individuals above a certain age or who have some risk factors for developing cognitive impairment, longitudinal performance data can be used with a high level of accuracy to determine which patients will require close monitoring. As treatments for Alzheimer’s Disease continue to develop, this ability to determine who will require close monitoring can allow more invasive and expensive tests to be reserved for them. This can also allow for early intervention, which can be crucial in treatment or prevention of the disease.

## Data Availability

All relevant data are available online.

https://www.kaggle.com/mukherji/plos-one-ad-project

